# Examining the association between male circumcision and the prevalence of self-reported sexually transmitted infections among adolescent boys and men (15-49 old) in Malawi, Rwanda, Zambia and Zimbabwe

**DOI:** 10.1101/2025.10.02.25337212

**Authors:** Jef Vanhamel, Tom Smekens, Wole Ameyan, Augustine Choko, Webster Mavhu, Mwelwa M Phiri, Bernadette Hensen

## Abstract

In 2007, the World Health Organization recommended voluntary medical male circumcision (VMMC) as an effective HIV prevention strategy in countries with a high HIV burden and low prevalence of male circumcision. While there is some evidence that VMMC lowers the risk of other sexually transmitted infections (STIs), its population-level impact beyond HIV remains uncertain. This study aimed to examine population and individual level associations between male circumcision (scale-up) and self-reported STIs (defined as a self-reported STI diagnosis or symptoms of a genital soar, ulcer, or discharge, in the past 12 months) among sexually active adolescent boys and men aged 15–49. We analysed nationally representative data from Demographic and Health Surveys conducted before (2004-2006) and after (2015-2020) VMMC scale-up in Malawi, Rwanda, Zambia, and Zimbabwe using a multilevel logistic regression approach adjusted for potential confounders. Our analysis included 41,094 respondents; circumcision coverage increased over time in all countries except Zimbabwe (11.9% before vs. 12.8% after VMMC scale-up; p=0.821). STI prevalence also increased, from 5.3% (95% CI 4.8-5.8) before to 8.2% (95% CI 7.7-8.7; p<0.001) after VMMC scale-up, with variations in the relative increase across countries. At population level, we found a higher odds of self-reported STIs after compared to before VMMC scale-up (aOR 1.52; 95% CI 1.33-1.74; p<0.001). We found no association between circumcision status and self-reported STIs at individual-level (aOR 1.03; 95% CI 0.92-1.17; p=0.55). These findings suggest that while VMMC protects against HIV and some STIs, its impact on reducing the STI burden at population level in Eastern and Southern Africa is limited. As STI rates remain high, especially among young men, our results underscore the need for comprehensive, male-oriented sexual health strategies. Reducing service delivery barriers, combined with integrating VMMC into person-centred sexual health services for men, may help address persistent gaps in STI prevention.

## Introduction

Three randomised controlled trials showed that medical male circumcision reduces the risk of heterosexual HIV acquisition among HIV negative men by 53-60% [1–3]. This evidence led the World Health Organization (WHO) and the Joint United Nations Programme on HIV/AIDS (UNAIDS) to recommend voluntary medical male circumcision (VMMC) as an essential component of a comprehensive HIV prevention strategy in 2007 [4]. In 2008, 14 countries in Eastern and Southern Africa with a high burden of HIV yet low prevalence of male circumcision were prioritised for VMMC programming (a fifteenth country was added in 2018), with a goal of 90% coverage among adolescent boys and men aged 15-49 by 2025 [5]. Between 2016–2020, approximately 18 million VMMC procedures were performed, far off the 2020 target of 25 million [6,7]. The WHO subsequently reinforced its recommendation on VMMC in 2020, and by the end of 2023 approximately 37.5 million VMMC procedures were performed in the prioritised countries [8,9].

Although VMMC programmes are promoted primarily for HIV prevention, evidence suggests that male circumcision offers partial protection against heterosexual transmission of other sexually transmitted infections (STIs), such as syphilis and herpes simplex virus type 2 (HSV-2) [10–12]. Biological mechanisms for this protective effect are not fully understood, but it is well established that the inner foreskin of the penis is highly susceptible to infections due to its warm and moist environment, and that its removal reduces infection susceptibility [1,13]. Moreover, the keratinisation of the glans penis (i.e. the penile head) when not protected by the foreskin results in fewer micro-abrasions during sexual contact, and hence reduced entry points for pathogens [1]. VMMC programmes do not focus solely on the surgical removal of the foreskin, but offer a comprehensive package of services, including HIV testing, sexual health promotion (including condom distribution), and screening and referrals for treatment of STIs, which may also have positive effects on reducing the STI burden [14]. In addition to benefits for circumcised men, epidemiological studies have demonstrated that female partners of medically circumcised men are at decreased risk of some STIs, including several oncogenic human papilloma virus (HPV) types [15,16].

It is unclear, however, whether these dynamics contribute to lower STI transmission at population level. This is in stark contrast with the evidence for the effectiveness of VMMC in reducing HIV incidence, which has been demonstrated in several observational studies in populations with diverse HIV risk and changing epidemic contexts [17]. The protective effect of VMMC on STI acquisition and transmission, on the other hand, has been documented predominantly in research settings, before VMMC scale-up, with effectiveness varying according to type of STI and several contextual factors (e.g. study setting and population) [18]. A recent review showed that there is convincing evidence for the protective effect of VMMC against acquisition and transmission of HSV-2, HPV and syphilis in heterosexual men and women [18]. However, the evidence for a protective effect of VMMC on the acquisition and transmission of *Chlamydia Trachomatis* (CT) and *Neisseria Gonorrhoea* (NG) is more limited and conflicting [18,19]. Moreover, it remains unclear whether scaling-up VMMC programmes has resulted in any measurable impact on the STI burden in countries prioritised for scale-up. No studies to our knowledge have focused on multi-country analyses using nationally representative data collected after VMMC scale-up to estimate the effect of male circumcision on STIs beyond HIV. Given the high and increasing STI rates observed globally, developing a better understanding of any protective effect of male circumcision on STIs other than HIV has important public health implications [20].

This study had two objectives: (1) to explore population level association between VMMC scale-up (i.e. increase in male circumcision coverage) and the prevalence of self-reported STIs, and (2) to examine individual level association between male circumcision status and self-reported STIs among sexually active adolescent boys and men (15-49 years), using data from the Demographic and Health Surveys (DHS) conducted in four countries prioritised for VMMC scale-up in Eastern and Southern Africa: Malawi, Rwanda, Zambia and Zimbabwe.

## Methods

### Study design

This paper is based on a secondary analysis of cross-sectional data from nationally representative population-based DHS surveys from four Eastern and Southern African countries prioritised for VMMC scale-up by the WHO. Only countries that had available datasets for both pre- and post-2008 time points, with the most recent DHS survey conducted in 2015-2020, and that had similar levels of VMMC coverage in the pre-2008 surveys, were included (see Table 1). The DHS methodology, including sampling strategy, is comprehensively detailed in the individual DHS survey country reports [21]. The surveys are designed to ensure that – when appropriately weighted – the study populations are representative at national and subnational level.

**Table 1.** DHS country surveys included in this study.

| Pre-2008 (pre-VMMC scale-up) surveys | Post-2008 (post-VMMC scale-up) surveys |
| --- | --- |
| Malawi (2004) | Malawi (2015-2016) |
| Rwanda (2005) | Rwanda (2019-2020) |
| Zambia (2007) | Zambia (2018) |
| Zimbabwe (2005-2006) | Zimbabwe (2015) |

### Study population and variables

We included adolescent boys and men (15-49 years) targeted for VMMC and who reported having ever had sex, and who responded to questions regarding their circumcision status and self-reported STIs in the surveys.

#### Main outcome variable

Self-reported STIs in the past 12 months; this was a composite variable we constructed by combining three variables, including “self-reported STI diagnosis in the past 12 months”, “self-reported symptoms of genital discharge in the past 12 months”, and “self-reported symptoms of a genital soar or ulcer in the past 12 months”. Participants responding ‘*yes*’ to any of these three questions were coded as a case of self-reported STI. Participants who answered ‘*no*’ to all three questions were coded as not a case of self-reported STI. Participants responding ‘*I don’t know*’ to any of the three questions while not responding ‘*yes*’ to the other two questions were dropped from the analysis.

#### Main exposure variable

We used two main exposure variables, corresponding to our two research objectives: (1) to explore whether temporal changes in VMMC implementation (i.e. VMMC scale-up) were associated with the outcome, we created a dichotomous variable “survey timing” as a population-level exposure variable. This variable indicated whether respondents participated in surveys conducted either before or after 2008; (2) we used self-reported circumcision status (i.e. circumcised or not circumcised) as main exposure variable for the individual level analysis. The post-2008 surveys allowed for differentiation of circumcision status according to who performed the circumcision (i.e. a medical professional or traditional practitioner), whereas the pre-2008 surveys did not collect this information. In the post-2008 surveys, we used this information to classify participants as “not circumcised”, “traditionally circumcised” or “medically circumcised”.

#### Other variables

We included socio-demographic and behavioural variables to be assessed for their association with the main exposure and outcome, as potential confounders. Age was categorised into five-year intervals (15–19, 20–24, etc.), and participants’ place of residence was classified as either rural or urban. Mobility status was defined based on whether a participant had spent more than one month away from home in the past year. Occupational status was measured by current employment (employed vs. not employed), and educational attainment was captured as the highest level of education completed. Religious affiliation varied by country, so original response categories were harmonised across surveys into three groups: Catholic/Orthodox, Protestant/Other Christian, and Muslim/Other/None. Marital status was reclassified into three categories: never married, divorced/widowed/separated, and married or living together. Socioeconomic status was represented by the wealth index quintile, a composite measure based on household ownership of selected assets and amenities. Exposure to mass media was evaluated through self-reported frequency of engaging with newspapers or magazines, radio, and television.

Sexual risk behaviour was captured using several indicators: age at sexual debut (classified as under 18 years, 18 years or older, or unknown), condom use during the last sexual encounter (yes or no), number of sexual partners in the past 12 months (none, one, two, or more than two), and whether the participant had paid for sex in the same period (yes or no). Participants’ knowledge of HIV and STIs was assessed using three statements: whether the risk of HIV can be reduced by having fewer sexual partners, whether it can be reduced through condom use, and whether a healthy-looking person can have HIV. Each statement had response options of ‘*yes*’, ‘*no’*, or ‘*don’t know’*.

Lastly, we included HIV serostatus as a potential confounding variable. This data was obtained from blood spots using a finger prick, accompanied by voluntary HIV counselling, as part of the country survey procedures. The detailed HIV testing procedures followed national guidelines and can be accessed in the separate country survey reports [21]. The barcodes identifying the HIV test results were linked with the individual men’s datasets.

### Data analysis

We used descriptive statistics to summarise the study population by country and per survey round. We used Chi-square statistics to compare characteristics between circumcised and uncircumcised adolescent boys and men across both surveys within each country. To address our two research objectives, we pooled all surveys into one dataset and conducted two separate multilevel logistic regression analyses adjusted for individual level confounders:

**(1) Before-after analysis of VMMC programme impact:** to examine whether VMMC scale-up was associated with changes in self-reported STIs over time, we assessed the association between self-reported STI status and “survey timing” (before vs. after 2008) as the main exposure variable at the population level. This analysis did not include individual circumcision status as a covariate, as the objective was to assess the impact of VMMC programme implementation.
**(2) Individual level association between circumcision status and self-reported STIs**: we used individual circumcision status (circumcised vs. not circumcised) as the main exposure variable. This analysis controlled for survey timing (before vs. after 2008) as a potential confounder.

For both logistic regression analyses, the same proximal-distal conceptual framework was developed to avoid over-adjusting for variables that may act as mediators (Figure 1). In this framework, if more proximal variables were found to be associated with the outcome, variables located more distally from this variable were adjusted for these variables but not vice versa. Variables located at the same level were adjusted for each other (e.g. sexual risk behaviour, circumcision and HIV status). For both analyses, we first conducted bivariate analyses between variables of interest and self-reported STIs. Then, we conducted multivariate analyses adjusted for potential individual level confounders following the framework. In the adjusted model, we only controlled for independent variables found to be associated with the outcome at the p<0.2 significance level in bivariate analyses. Measures of association were expressed as odds ratios (OR) with 95% confidence intervals (CI) and their p-values. Measures of variation between the groups were expressed as the variance estimate of the random slopes in the null model (without any explanatory variables) and in the multivariate multilevel model.

**Figure 1.**
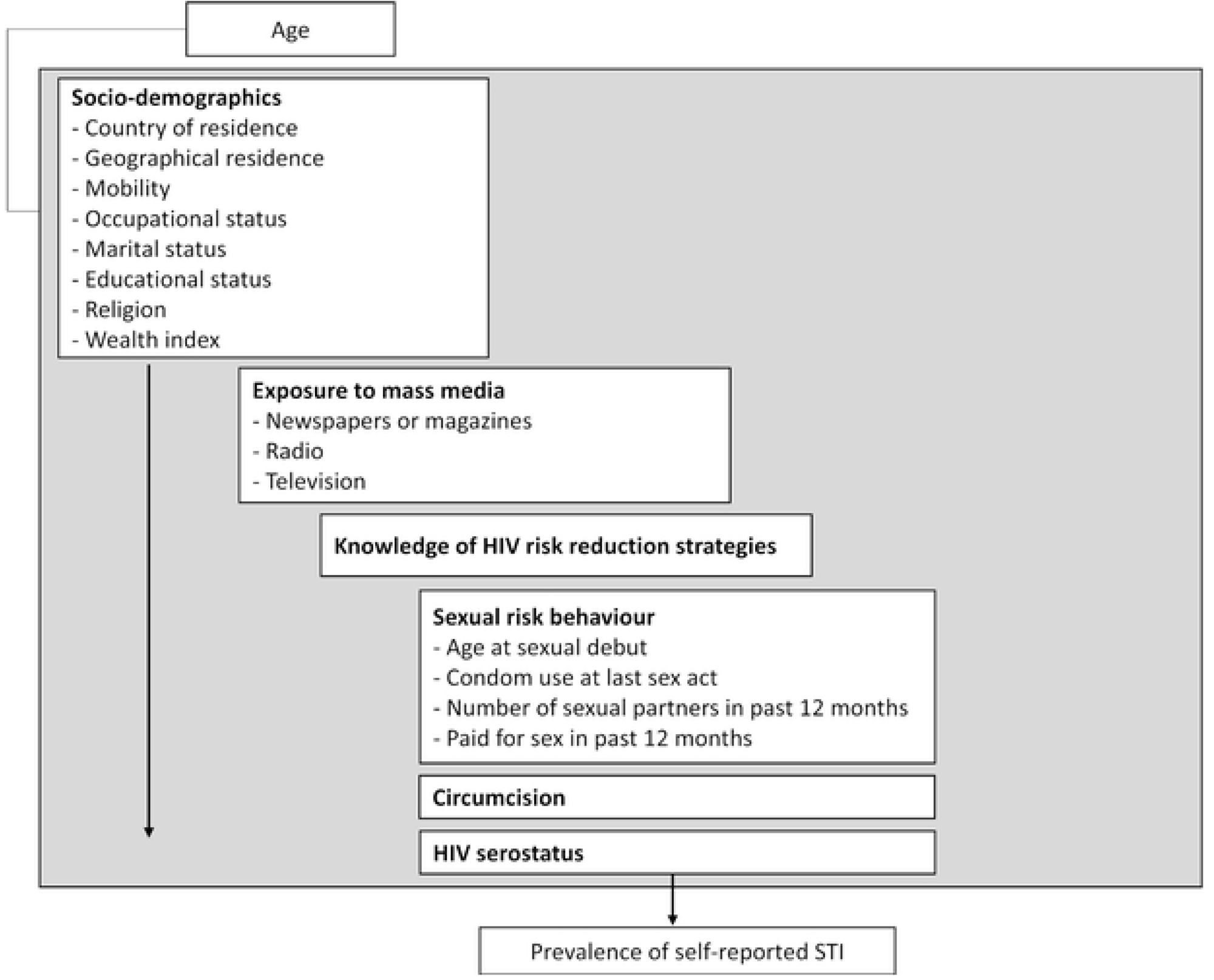
Conceptual framework for exploring the association between variables of interest and self-reported STIs.

We conducted two sensitivity analyses using the same multilevel analysis approach described above:

1. **STI diagnosis-only analysis**: using the pooled dataset, with the same covariates as in the main analysis, but restricting the outcome to self-reported STI diagnoses only (excluding STI symptoms).
2. **Medical circumcision analysis**: using only surveys conducted after 2008 (that distinguished between medical and traditional circumcision), maintaining the full STI outcome definition (including symptoms), but excluding individuals who reported being traditionally circumcised to focus specifically on medical circumcision effects.

We adjusted for sampling weights, clustering and stratification, as per the Guide to DHS Statistics, to derive proportions in descriptive statistics and in all bivariate and multivariate analyses, to ensure representativeness [22]. Multilevel models were fitted using the “glmer” function. All analyses were carried out in R (version 4.4.0) [23].

### Ethics statement

Procedures and questionnaires for standard DHS surveys have been reviewed and approved by the ICF Institutional Review Board (IRB). Additionally, country-specific DHS survey protocols are reviewed by the ICF IRB and typically by an IRB in the host country. ICF IRB ensures that the survey complies with the U.S. Department of Health and Human Services regulations for the protection of human subjects (45 CFR 46), while the host country IRB ensures that the survey complies with laws and norms of the nation. Both verbal and written informed consent were obtained from the respondents before their voluntary participation. Written informed consent was obtained from the parent/guardian of each participant under 18 years of age. Given that this current study involved a secondary analysis of DHS data, we did not require any further ethics approval. The first author received access to the data from the DHS programme on 6 June 2024. The authors did not have access to information that could identify individual participants during or after data collection.

## Results

Our study included a total of 41,094 adolescent boys and men aged 15-49, 15,660 of whom were included in the pre-2008 survey round and 25,434 in the post-2008 survey round (see Figure 2).

**Figure 2.**
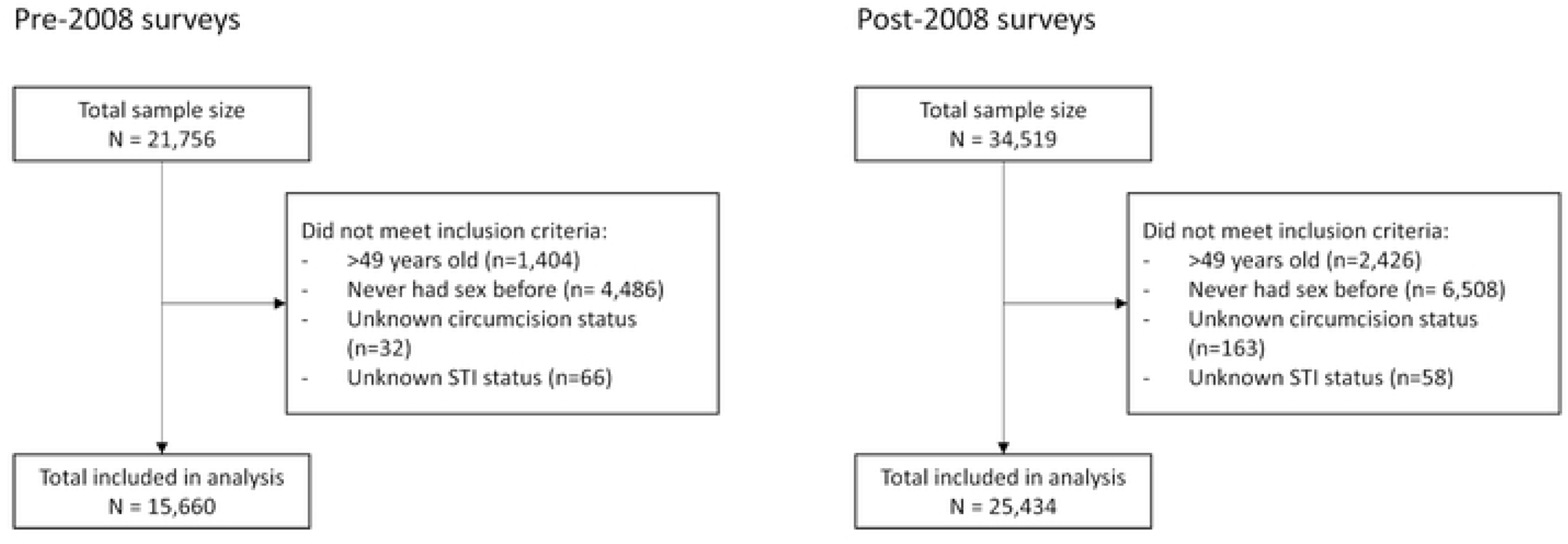
Selection of the final study population for each survey round included in the analysis.

Most respondents were between 20 and 39 years (70.2% in the pre- and 66.8% post-2008 surveys; Table 2 and 3). In both survey rounds, most respondents were not circumcised (85.9% pre-2008 and 71.1% post-2008). Circumcised boys and men in both survey rounds were more likely than uncircumcised boys and men to be based in urban areas (37.1% vs. 33.0%, respectively, in pre-2008 surveys; p=0.013; 40.0% vs. 29.0%, respectively, in post-2008 surveys; p<0.001), be of non-Christian religion (28.7% vs. 14.6%, respectively, in pre-2008 surveys; p<0.001; 14.0% vs. 9.0%, respectively, in post-2008 surveys; p<0.001), belong to the richest quintile (27.8% vs. 22.6%, respectively, in pre-2008 surveys; p=0.001; 31.8% vs. 19.9%, respectively, in post-2008 surveys; p<0.001) and have an age below 18 years at sexual debut (48.2% vs. 41.7%, respectively, in pre-2008 surveys; p<0.001; 45.1% vs. 40.5%, respectively, in post-2008 surveys; p<0.001). The prevalence of self-reported STIs did not differ significantly between circumcised and non-circumcised respondents in both survey rounds. Supplementary files S1 and S2 show the characteristics for each country separately.

**Table 2.** Socio-demographics and sexual risk behaviour by circumcision status among men aged 15-49 in the pooled pre-2008 population-based surveys.

|  | **Total**<br>**N (%)** | **Not circumcised**<br>**N (%) \*** | **Circumcised**<br>**N (%) \*** | **P-value<sup>§</sup>** |
| --- | --- | --- | --- | --- |
| **Total** | 15660 (100) | 13143 (85.9) | 2517 (14.1) |  |
| **Age group** |  |  |  |  |
| 15-19 | 1761 (10.9) | 1450 (10.7) | 311 (11.6) | 0.757 |
| 20-24 | 3056 (19.5) | 2579 (19.6) | 477 (18.6) |  |
| 25-29 | 3073 (19.8) | 2594 (19.8) | 479 (20.0) |  |
| 30-34 | 2745 (17.6) | 2294 (17.6) | 451 (18.1) |  |
| 35-39 | 2066 (13.3) | 1741 (13.4) | 325 (12.9) |  |
| 40-44 | 1596 (10.3) | 1333 (10.2) | 263 (10.6) |  |
| 45-49 | 1363 (8.6) | 1152 (8.6) | 211 (8.1) |  |
| **Country of residence** |  |  |  |  |
| Malawi | 2608 (16.6) | 1944 (15.0) | 664 (26.4) | <0.001 |
| Rwanda | 3025 (19.3) | 2634 (20.0) | 391 (15.4) |  |
| Zambia | 5075 (32.0) | 4200 (32.1) | 875 (31.2) |  |
| Zimbabwe | 4952 (32.1) | 4365 (33.0) | 587 (27.0) |  |
| **Geographic residence** |  |  |  |  |
| Urban | 5063 (33.6) | 4180(33.0) | 883 (37.1) | 0.013 |
| Rural | 10597 (66.4) | 8963 (67.0) | 1634 (62.9) |  |
| **Mobility (away from home >1 month)** |  |  |  |  |
| No | 4675 (65.7) | 3943 (65.9) | 732 (64.5) | 0.553 |
| Yes | 2426 (33.9) | 1999 (33.8) | 427 (34.8) |  |
| **Current occupational status** |  |  |  |  |
| Unemployed | 4396 (28.0) | 3685 (28.1) | 711 (27.3) | 0.786 |
| Employed | 11226 (71.9) | 9427 (71.7) | 1799 (72.6) |  |
| **Education status** |  |  |  |  |
| No education | 1203 (7.9) | 1154 (8.0) | 49 (7.3) | 0.606 |
| Primary level | 7363 (46.4) | 6988 (46.4) | 375 (46.1) |  |
| Secondary level | 6238 (40.1) | 5861 (40.0) | 377 (40.3) |  |
| Higher level | 856 (5.7) | 827 (5.6) | 29 (6.3) |  |
| **Religion** |  |  |  |  |
| Catholic / Orthodox | 3573 (23.5) | 3151 (24.5) | 422 (17.5) | <0.001 |
| Protestant / Other Christian | 9440 (59.9) | 8051 (60.8) | 1389 (53.8) |  |
| Muslim/ None / Other | 2646 (16.6) | 1940 (14.6) | 706 (28.7) |  |
| **Marital status** |  |  |  |  |
| Never married | 4686 (29.6) | 3919 (29.6) | 767 (30.0) | 0.517 |
| Divorced / Separated / Widowed | 794 (5.0) | 657 (4.9) | 137 (5.5) |  |
| Married or living together | 10180 (65.4) | 8567 (65.5) | 1613 (64.5) |  |
| **Wealth index** |  |  |  |  |
| Poorest | 2689 (17.1) | 2329 (17.6) | 360 (14.0) | 0.001 |
| Poorer | 2729 (16.9) | 2275 (17.0) | 454 (16.5) |  |
| Middle | 3035 (18.3) | 2531 (18.3) | 504 (18.2) |  |
| Richer | 3821 (24.4) | 3241 (24.5) | 580 (23.6) |  |
| Richest | 3386 (23.3) | 2767 (22.6) | 619 (27.8) |  |
| **Exposure to mass media** |  |  |  |  |
| **Newspaper or magazines** |  |  |  |  |
| Not at all | 7723 (49.3) | 6565 (50.0) | 1158 (45.2) | <0.001 |
| Less than once a week | 3383 (21.3) | 2852 (21.3) | 531 (21.8) |  |
| At least once a week | 4531 (29.3) | 3710 (28.7) | 821 (32.9) |  |
| **Radio** |  |  |  |  |
| Not at all | 2202 (13.3) | 1869 (13.4) | 333 (12.6) | 0.598 |
| Less than once a week | 1849 (11.7) | 1577 (11.8) | 272 (10.9) |  |
| At least once a week | 11584 (74.9) | 9676 (74.7) | 1908 (76.3) |  |
| **Television** |  |  |  |  |
| Not at all | 8865 (55.3) | 7475 (55.7) | 1390 (53.1) | 0.091 |
| Less than once a week | 2151 (13.9) | 1785 (13.6) | 366 (15.5) |  |
| At least once a week | 4627 (30.7) | 3870 (30.6) | 757 (31.4) |  |
| **HIV knowledge** |  |  |  |  |
| **Can reduce HIV risk by fewer partners (baseline = those aware of HIV or AIDS)** |  |  |  |  |
| No | 1684 (11.5) | 1408 (11.4) | 276 (12.0) | 0.127 |
| Yes | 13755 (87.6) | 11567 (87.7) | 2188 (86.6) |  |
| Don't know | 149 (0.01) | 115 (0.01) | 34 (1.4) |  |
| **Can reduce HIV risk by condom use (baseline = those aware of HIV or AIDS)** |  |  |  |  |
| No | 2504 (16.2) | 2110 (16.2) | 394 (16.0) | 0.131 |
| Yes | 12748 (81.6) | 10684 (81.5) | 2064 (82.5) |  |
| Don't know | 321 (2.1) | 283 (2.2) | 38 (<0.01) |  |
| **Healthy looking people can have HIV (baseline = those aware of HIV or AIDS)** |  |  |  |  |
| No | 1049 (6.6) | 882 (6.6) | 167 (6.3) | 0.284 |
| Yes | 14391 (92.3) | 12080 (92.3) | 2311 (92.2) |  |
| Don't know | 173 (1.1) | 138 (1.0) | 35 (<0.01) |  |
| **Sexual risk behaviour** |  |  |  |  |
| **Age at sexual debut** |  |  |  |  |
| <18 | 6865 (42.6) | 5591 (41.7) | 1274 (48.2) | <0.001 |
| ≥18 | 8536 (55.8) | 7336 (56.7) | 1200 (50.1) |  |
| Don't know | 259 (1.6) | 216 (1.6) | 43 (1.7) |  |
| **Condom use at last sex act (baseline = at least 1 sexual partner in past 12 months)** |  |  |  |  |
| No | 10739 (80.4) | 8989 (80.4) | 1750 (80.6) | 0.670 |
| Yes | 2678 (19.6) | 2242 (19.6) | 436 (19.4) |  |
| Don't know | 5 (<0.01) | 5 (<0.01) | 0 |  |

| Number of sexual partners in past 12 months |  |  |  |  |
| --- | --- | --- | --- | --- |
| 0 | 2228 (14.5) | 1897 (14.5) | 331 (14.0) | 0.311 |
| 1 | 11527 (73.7) | 9688 (73.8) | 1839 (72.8) |  |
| 2 | 1615 (10.1) | 1319 (9.9) | 296 (11.4) |  |
| >2 | 265 (1.7) | 218 (1.7) | 47 (1.8) |  |
| Don't know | 12 (<0.01) | 11 (<0.01) | 1 (<0.01) |  |
| Paid for sex in the past 12 months |  |  |  |  |
| No | 13816 (93.3) | 11750 (94.4) | 2066 (86.5) | <0.001 |
| Yes | 987 (6.4) | 683 (5.4) | 304 (12.8) |  |
| HIV serostatus |  |  |  |  |
| Negative | 11190 (87.5) | 9431 (87.7) | 1759 (86.8) | 0.409 |
| Positive | 1584 (12.5) | 1345 (12.3) | 239 (13.2) |  |
| Self-reported STI in the last 12 months |  |  |  |  |
| No | 14830 (94.7) | 12447 (94.8) | 2383 (94.4) | 0.510 |
| Yes | 830 (5.3) | 696 (5.2) | 134 (5.6) |  |

\*Column percentages.
§ P-value of Chi-square test.

**Table 3.**
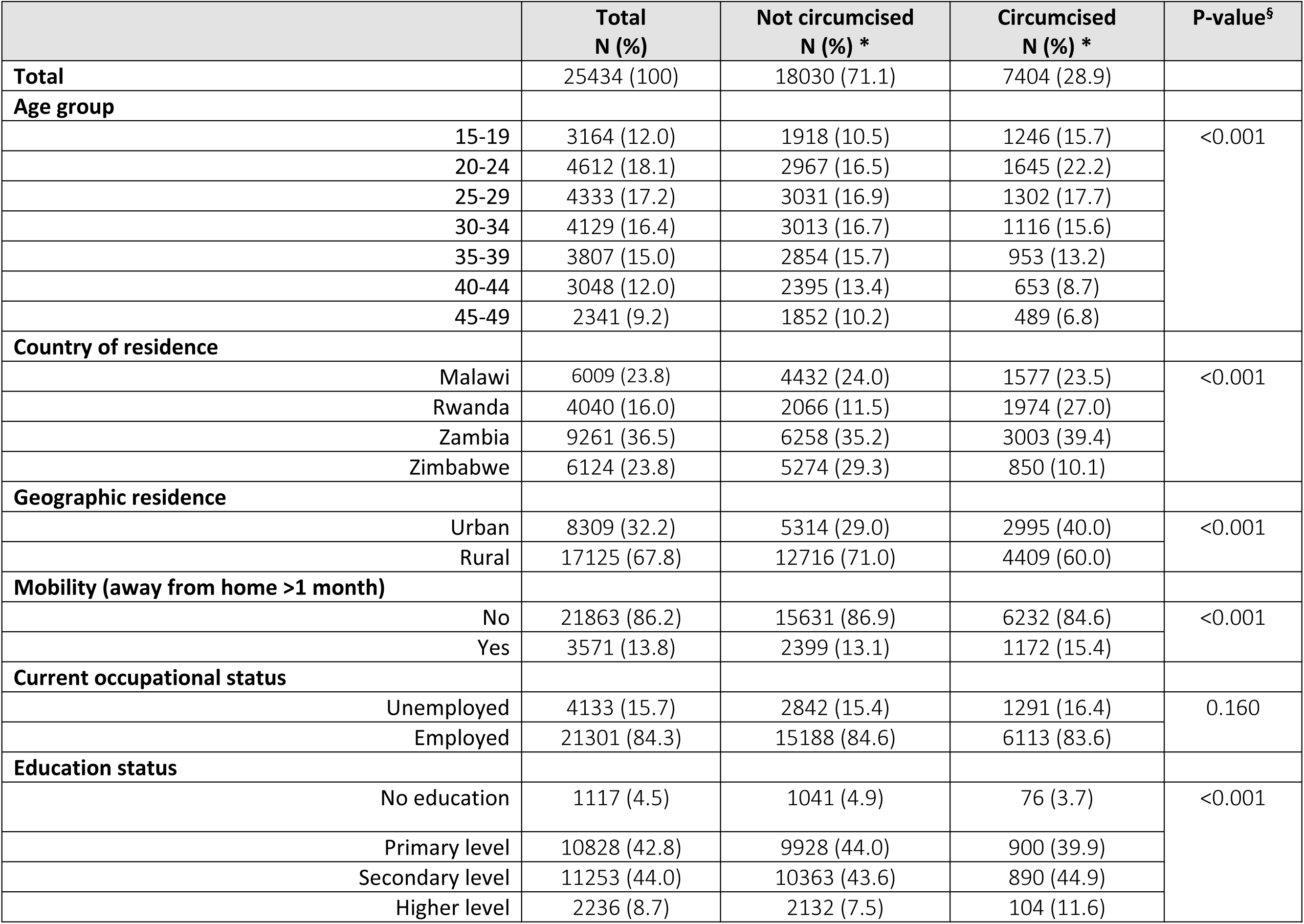
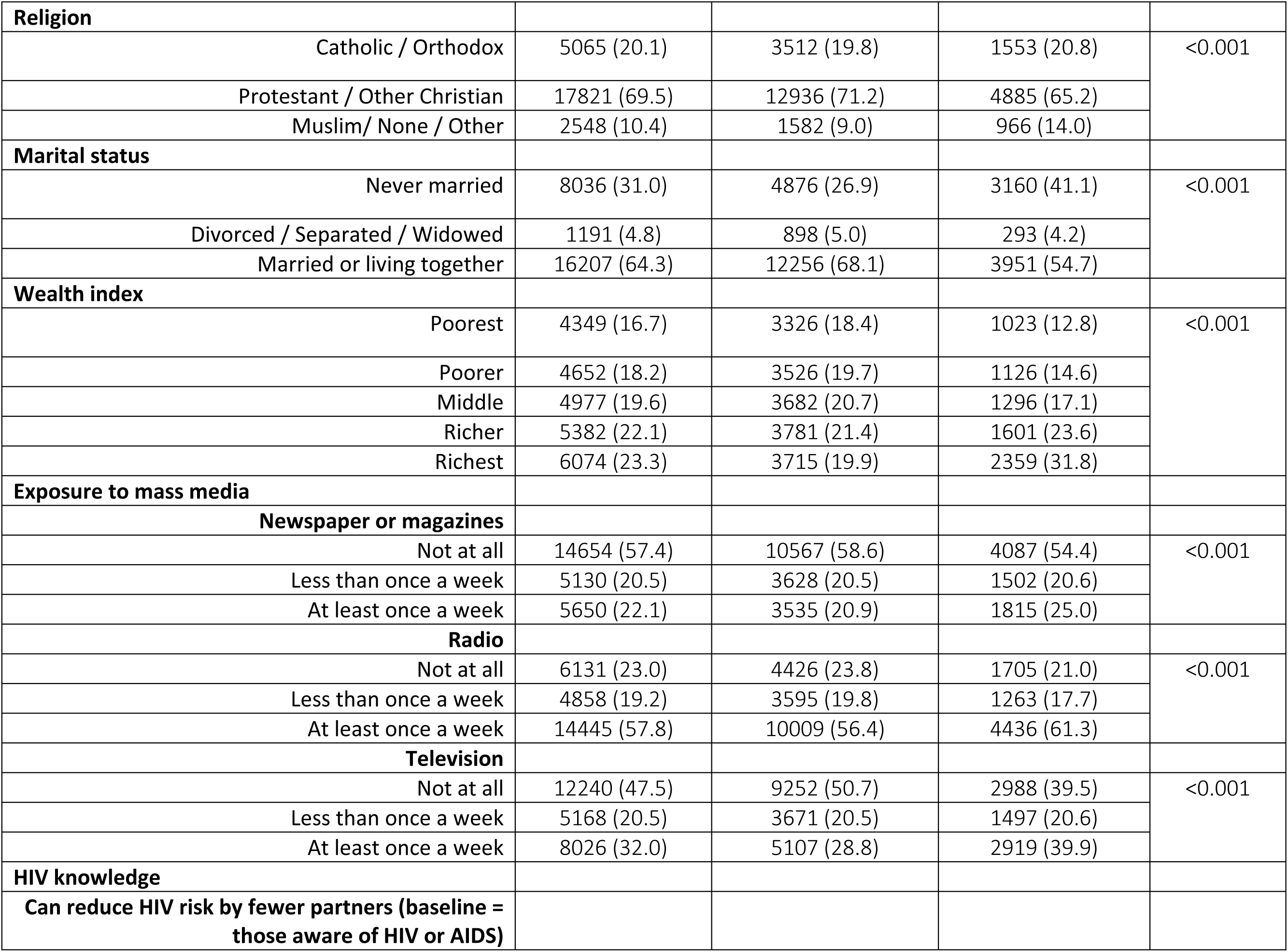
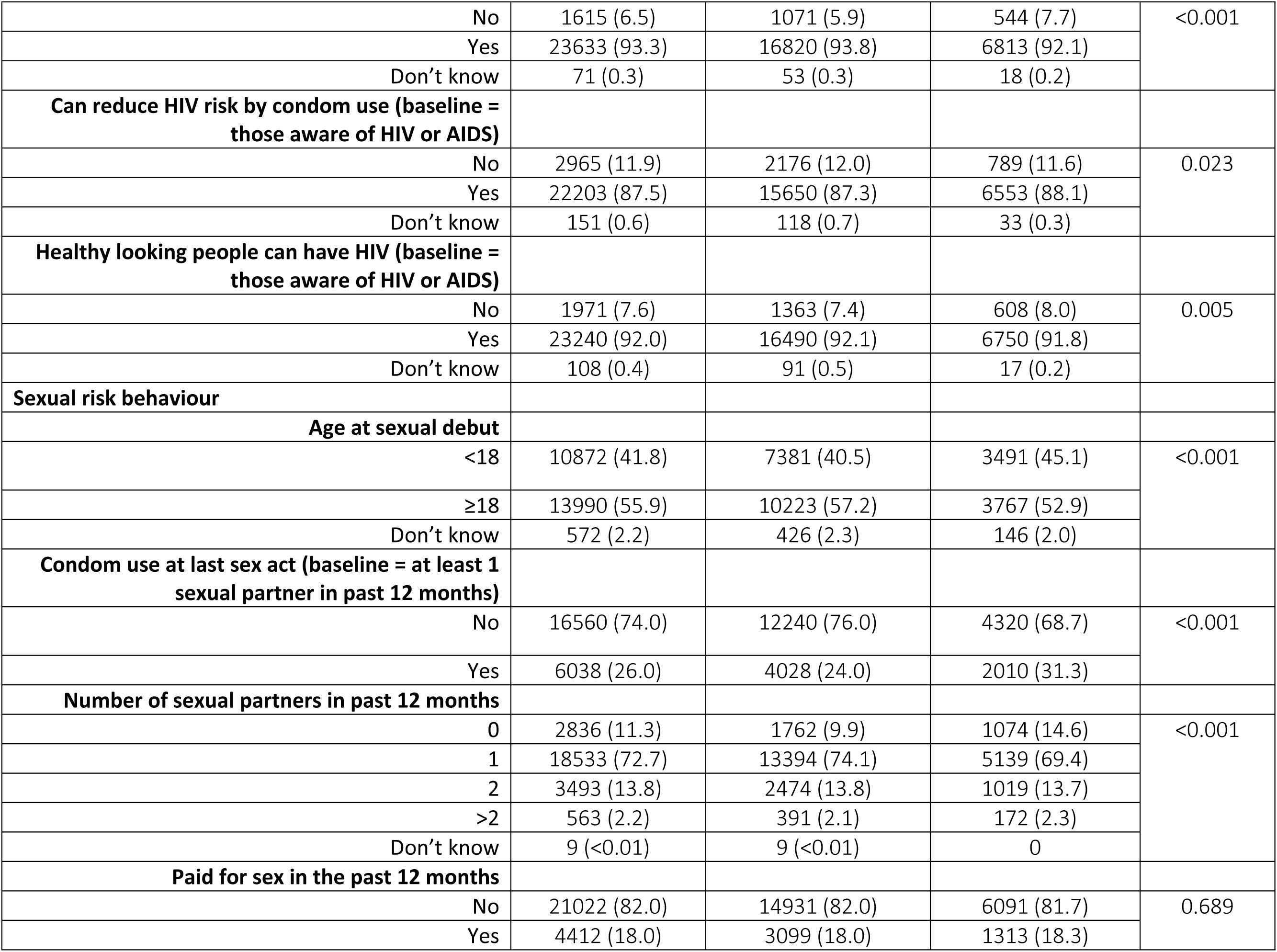
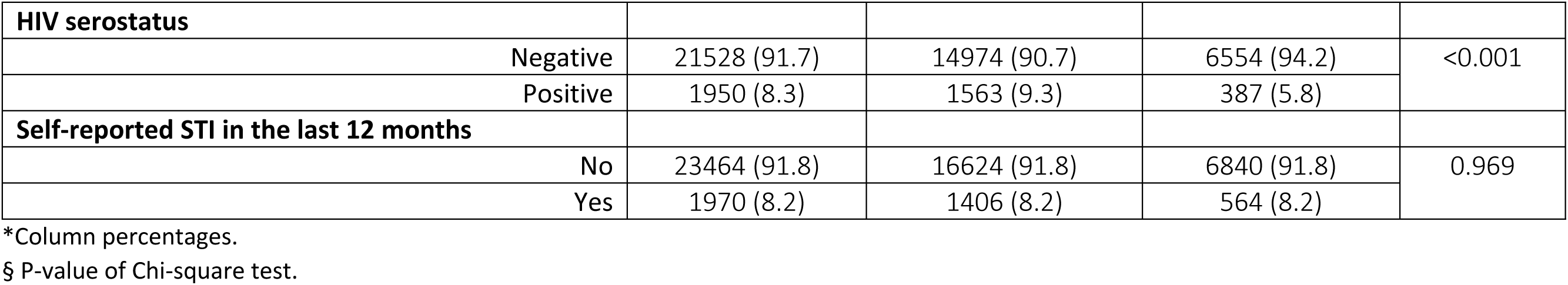
Socio-demographics and sexual risk behaviour by circumcision status among men aged 15-49 in the pooled post-2008 population-based surveys.

|  | **Total**<br>**N (%)** | **Not circumcised**<br>**N (%) \*** | **Circumcised**<br>**N (%) \*** | **P-value<sup>s</sup>** |
| --- | --- | --- | --- | --- |
| **Total** | 25434 (100) | 18030 (71.1) | 7404 (28.9) |  |
| **Age group** |  |  |  |  |
| 15-19 | 3164 (12.0) | 1918 (10.5) | 1246 (15.7) | <0.001 |
| 20-24 | 4612 (18.1) | 2967 (16.5) | 1645 (22.2) |  |
| 25-29 | 4333 (17.2) | 3031 (16.9) | 1302 (17.7) |  |
| 30-34 | 4129 (16.4) | 3013 (16.7) | 1116 (15.6) |  |
| 35-39 | 3807 (15.0) | 2854 (15.7) | 953 (13.2) |  |
| 40-44 | 3048 (12.0) | 2395 (13.4) | 653 (8.7) |  |
| 45-49 | 2341 (9.2) | 1852 (10.2) | 489 (6.8) |  |
| **Country of residence** |  |  |  |  |
| Malawi | 6009 (23.8) | 4432 (24.0) | 1577 (23.5) | <0.001 |
| Rwanda | 4040 (16.0) | 2066 (11.5) | 1974 (27.0) |  |
| Zambia | 9261 (36.5) | 6258 (35.2) | 3003 (39.4) |  |
| Zimbabwe | 6124 (23.8) | 5274 (29.3) | 850 (10.1) |  |
| **Geographic residence** |  |  |  |  |
| Urban | 8309 (32.2) | 5314 (29.0) | 2995 (40.0) | <0.001 |
| Rural | 17125 (67.8) | 12716 (71.0) | 4409 (60.0) |  |
| **Mobility (away from home >1 month)** |  |  |  |  |
| No | 21863 (86.2) | 15631 (86.9) | 6232 (84.6) | <0.001 |
| Yes | 3571 (13.8) | 2399 (13.1) | 1172 (15.4) |  |
| **Current occupational status** |  |  |  |  |
| Unemployed | 4133 (15.7) | 2842 (15.4) | 1291 (16.4) | 0.160 |
| Employed | 21301 (84.3) | 15188 (84.6) | 6113 (83.6) |  |
| **Education status** |  |  |  |  |
| No education | 1117 (4.5) | 1041 (4.9) | 76 (3.7) | <0.001 |
| Primary level | 10828 (42.8) | 9928 (44.0) | 900 (39.9) |  |
| Secondary level | 11253 (44.0) | 10363 (43.6) | 890 (44.9) |  |
| Higher level | 2236 (8.7) | 2132 (7.5) | 104 (11.6) |  |
| **Religion** |  |  |  |  |
| Catholic / Orthodox | 5065 (20.1) | 3512 (19.8) | 1553 (20.8) | <0.001 |
| Protestant / Other Christian | 17821 (69.5) | 12936 (71.2) | 4885 (65.2) |  |
| Muslim/ None / Other | 2548 (10.4) | 1582 (9.0) | 966 (14.0) |  |
| **Marital status** |  |  |  |  |
| Never married | 8036 (31.0) | 4876 (26.9) | 3160 (41.1) | <0.001 |
| Divorced / Separated / Widowed | 1191 (4.8) | 898 (5.0) | 293 (4.2) |  |
| Married or living together | 16207 (64.3) | 12256 (68.1) | 3951 (54.7) |  |
| **Wealth index** |  |  |  |  |
| Poorest | 4349 (16.7) | 3326 (18.4) | 1023 (12.8) | <0.001 |
| Poorer | 4652 (18.2) | 3526 (19.7) | 1126 (14.6) |  |
| Middle | 4977 (19.6) | 3682 (20.7) | 1296 (17.1) |  |
| Richer | 5382 (22.1) | 3781 (21.4) | 1601 (23.6) |  |
| Richest | 6074 (23.3) | 3715 (19.9) | 2359 (31.8) |  |
| **Exposure to mass media** |  |  |  |  |
| **Newspaper or magazines** |  |  |  |  |
| Not at all | 14654 (57.4) | 10567 (58.6) | 4087 (54.4) | <0.001 |
| Less than once a week | 5130 (20.5) | 3628 (20.5) | 1502 (20.6) |  |
| At least once a week | 5650 (22.1) | 3535 (20.9) | 1815 (25.0) |  |
| **Radio** |  |  |  |  |
| Not at all | 6131 (23.0) | 4426 (23.8) | 1705 (21.0) | <0.001 |
| Less than once a week | 4858 (19.2) | 3595 (19.8) | 1263 (17.7) |  |
| At least once a week | 14445 (57.8) | 10009 (56.4) | 4436 (61.3) |  |
| **Television** |  |  |  |  |
| Not at all | 12240 (47.5) | 9252 (50.7) | 2988 (39.5) | <0.001 |
| Less than once a week | 5168 (20.5) | 3671 (20.5) | 1497 (20.6) |  |
| At least once a week | 8026 (32.0) | 5107 (28.8) | 2919 (39.9) |  |
| **HIV knowledge** |  |  |  |  |
| **Can reduce HIV risk by fewer partners (baseline = those aware of HIV or AIDS)** |  |  |  |  |
| No | 1615 (6.5) | 1071 (5.9) | 544 (7.7) | <0.001 |
| Yes | 23633 (93.3) | 16820 (93.8) | 6813 (92.1) |  |
| Don't know | 71 (0.3) | 53 (0.3) | 18 (0.2) |  |
| Can reduce HIV risk by condom use (baseline = those aware of HIV or AIDS) |  |  |  |  |
| No | 2965 (11.9) | 2176 (12.0) | 789 (11.6) | 0.023 |
| Yes | 22203 (87.5) | 15650 (87.3) | 6553 (88.1) |  |
| Don't know | 151 (0.6) | 118 (0.7) | 33 (0.3) |  |
| Healthy looking people can have HIV (baseline = those aware of HIV or AIDS) |  |  |  |  |
| No | 1971 (7.6) | 1363 (7.4) | 608 (8.0) | 0.005 |
| Yes | 23240 (92.0) | 16490 (92.1) | 6750 (91.8) |  |
| Don't know | 108 (0.4) | 91 (0.5) | 17 (0.2) |  |
| Sexual risk behaviour |  |  |  |  |
| Age at sexual debut |  |  |  |  |
| <18 | 10872 (41.8) | 7381 (40.5) | 3491 (45.1) | <0.001 |
| ≥18 | 13990 (55.9) | 10223 (57.2) | 3767 (52.9) |  |
| Don't know | 572 (2.2) | 426 (2.3) | 146 (2.0) |  |
| Condom use at last sex act (baseline = at least 1 sexual partner in past 12 months) |  |  |  |  |
| No | 16560 (74.0) | 12240 (76.0) | 4320 (68.7) | <0.001 |
| Yes | 6038 (26.0) | 4028 (24.0) | 2010 (31.3) |  |
| Number of sexual partners in past 12 months |  |  |  |  |
| 0 | 2836 (11.3) | 1762 (9.9) | 1074 (14.6) | <0.001 |
| 1 | 18533 (72.7) | 13394 (74.1) | 5139 (69.4) |  |
| 2 | 3493 (13.8) | 2474 (13.8) | 1019 (13.7) |  |
| >2 | 563 (2.2) | 391 (2.1) | 172 (2.3) |  |
| Don't know | 9 (<0.01) | 9 (<0.01) | 0 |  |
| Paid for sex in the past 12 months |  |  |  |  |
| No | 21022 (82.0) | 14931 (82.0) | 6091 (81.7) | 0.689 |
| Yes | 4412 (18.0) | 3099 (18.0) | 1313 (18.3) |  |
| **HIV serostatus** |  |  |  |  |
| Negative | 21528 (91.7) | 14974 (90.7) | 6554 (94.2) | <0.001 |
| Positive | 1950 (8.3) | 1563 (9.3) | 387 (5.8) |  |
| **Self-reported STI in the last 12 months** |  |  |  |  |
| No | 23464 (91.8) | 16624 (91.8) | 6840 (91.8) | 0.969 |
| Yes | 1970 (8.2) | 1406 (8.2) | 564 (8.2) |  |

\*Column percentages.
§ P-value of Chi-square test.

Circumcision coverage and increases in circumcision coverage over time differed between countries (see Figure 3). The largest increase in circumcision coverage was observed in Rwanda, with the circumcision coverage among respondents increasing from 11.3% (95% CI 9.8 – 12.7%) in 2005 to 49.0% (95% CI 46.8 – 50.7%; p<0.001) in 2019-2020. The smallest – statistically insignificant – increase in circumcision coverage occurred in Zimbabwe, with 11.9% (95% CI 10.5 – 13.2%) of respondents circumcised in 2005-2006 compared to 12.8% (95% CI 11.1 – 13.6%; p=0.341) in 2015. The overall prevalence of self-reported STIs increased significantly from 5.3% (95% CI 4.8 – 5.8%) in the pre-2008 survey round, to 8.2% (95% CI 7.7 – 8.7%; p<0.001) in the post-2008 survey round. The prevalence of self-reported STIs also varied between countries and survey rounds, with the sharpest increase observed in Malawi, from 2.7% (95% CI 2.0 – 3.4%) in 2004 to 9.9% (95% CI 8.8 – 11.0%; p<0.001) in 2015-2016, and more modest increases in Zambia and Zimbabwe (see Figure 3).

**Figure 3.**
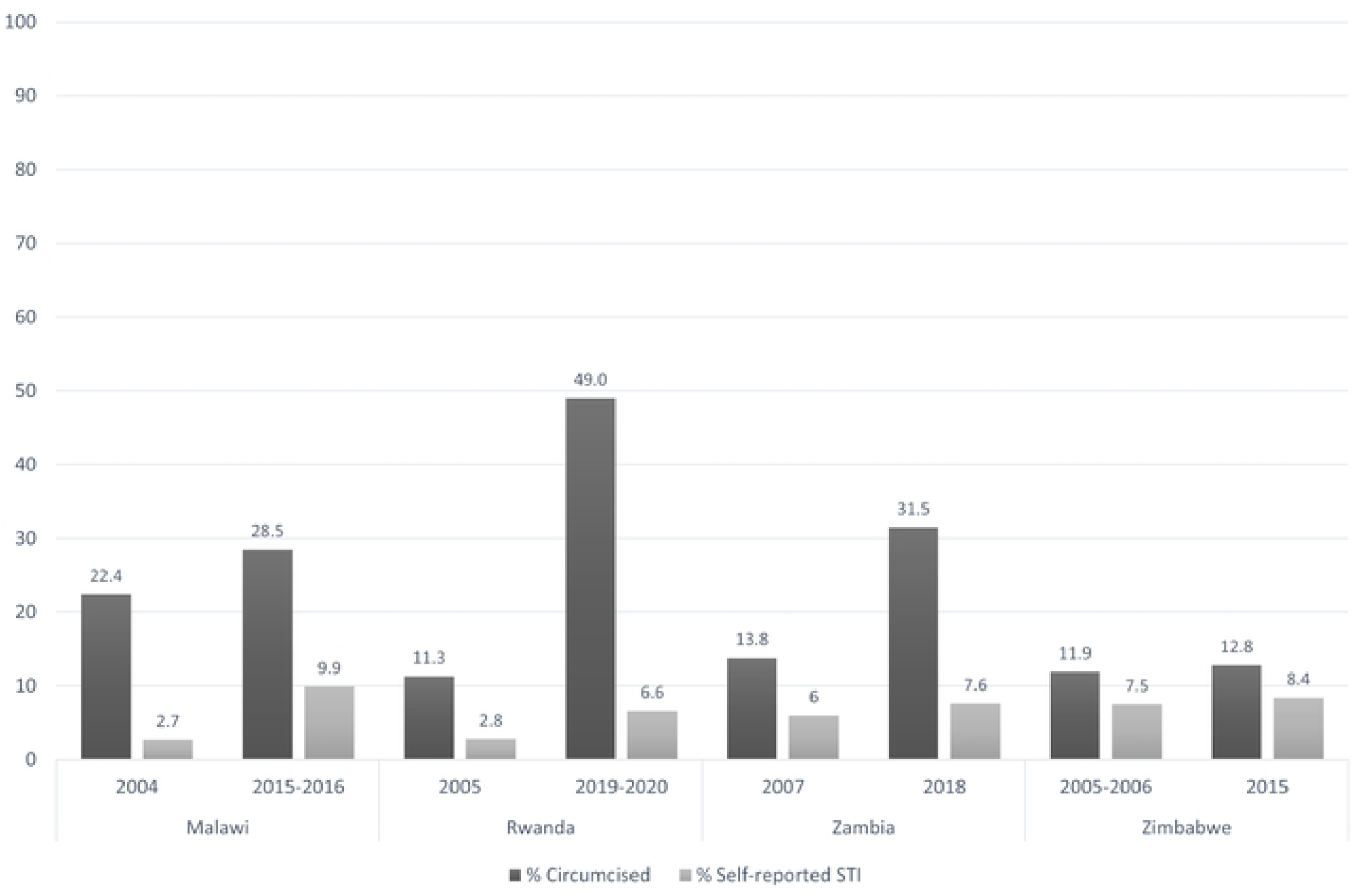
Percentage of men aged 15-49 that are circumcised (dark grey) and self-reported an STI (light grey) for each survey included in the analysis.

In the crude analyses, we found a higher odds of self-reported STIs in surveys conducted after 2008 (crude OR = 1.67; 95% CI 1.50 – 1.86; p<0.001; Table 4). After controlling for individual level confounders (but not circumcision), the OR decreased to 1.52 (95% CI 1.33 – 1.74; p<0.001). We found no significant association between individual level circumcision status and self-reported STIs (aOR = 1.03; 95% CI 0.92 – 1.17; p=0.547). Higher age at sexual debut was significantly associated with lower self-reported STIs (aOR = 0.76; 95% CI 0.70 – 0.83; p<0.001), whereas the odds of self-reported STIs increased with the number of sexual partners reported in the past 12 months (aOR for > 2 sexual partners = 5.66; 95% CI 4.41 – 7.27; p<0.001). Having paid for sex in the past 12 months was associated with a higher prevalence of self-reported STIs (aOR = 2.10; 95% CI 1.89 – 2.35; p<0.001), as was testing positive on the HIV test offered as part of the survey (aOR = 2.39; 95% CI 2.10 – 2.71; p<0.001).

**Table 4.**
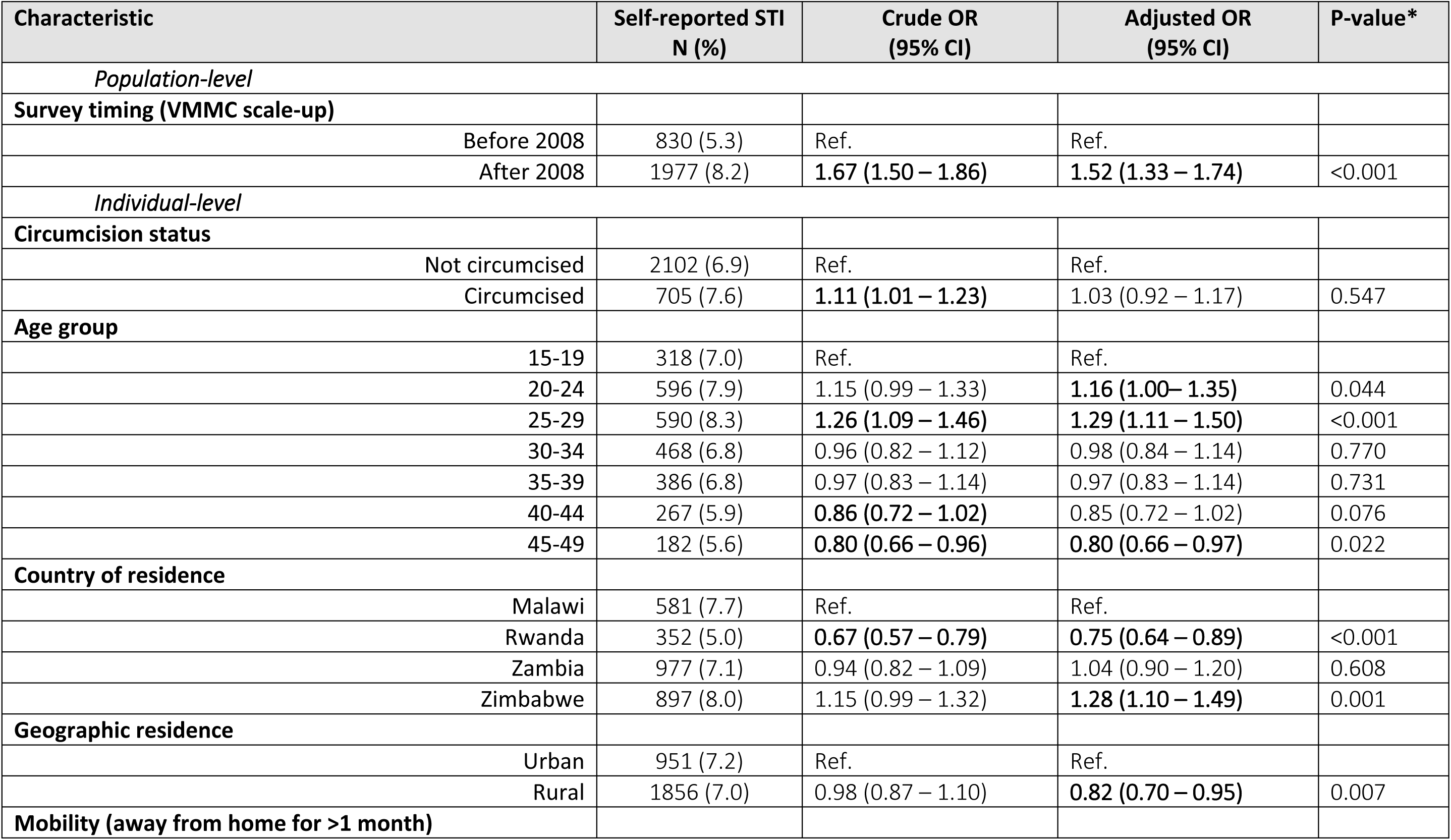
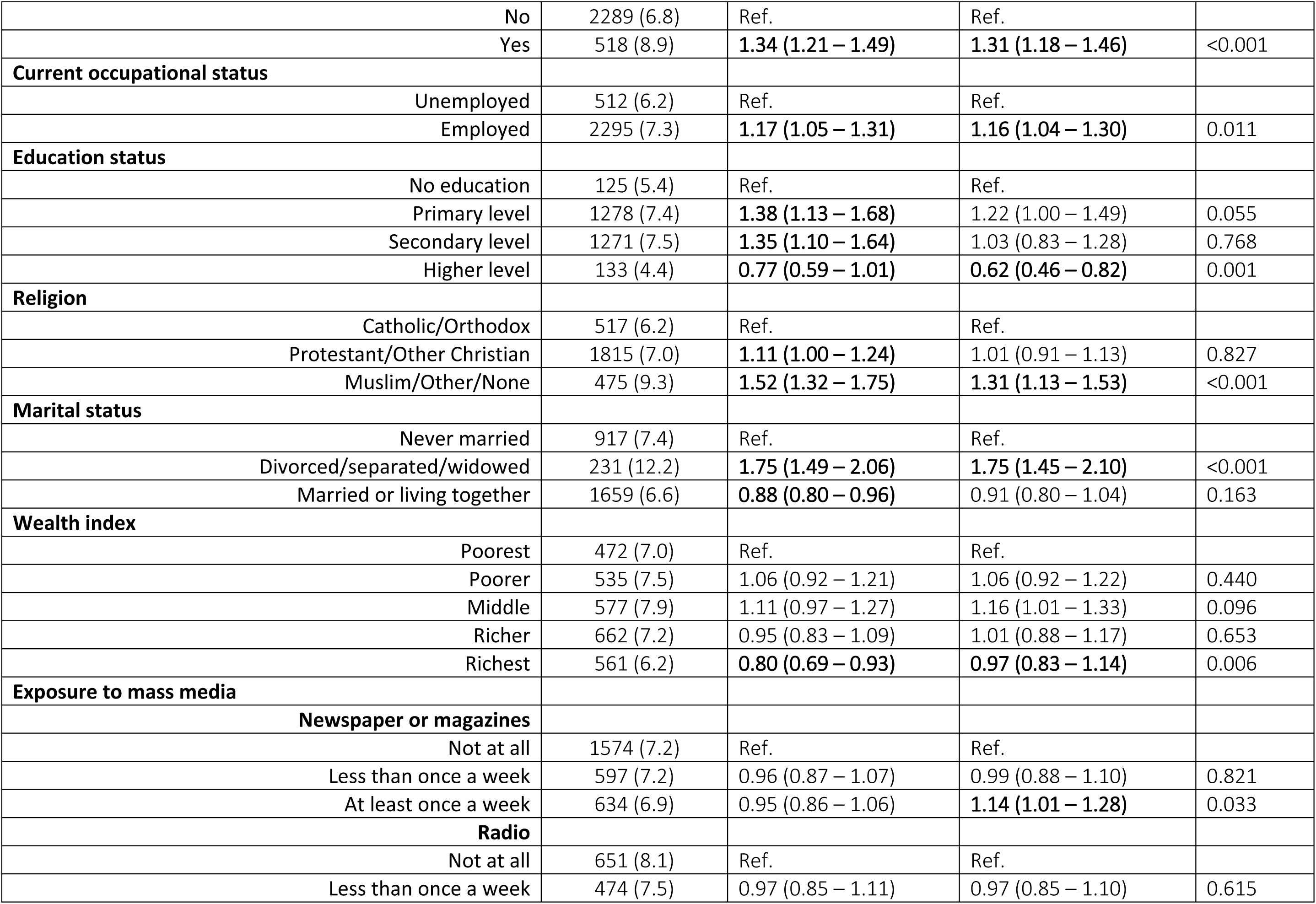
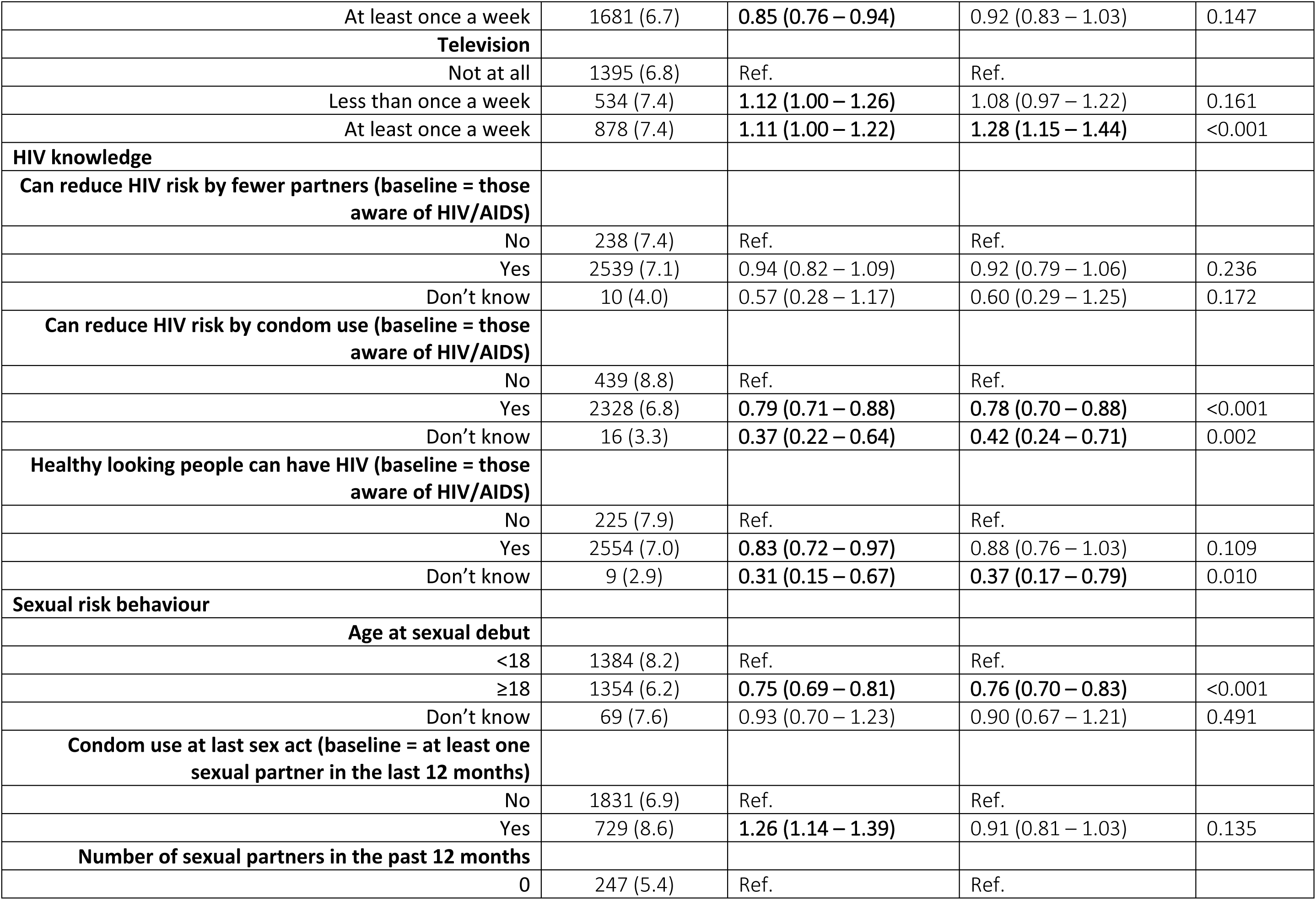
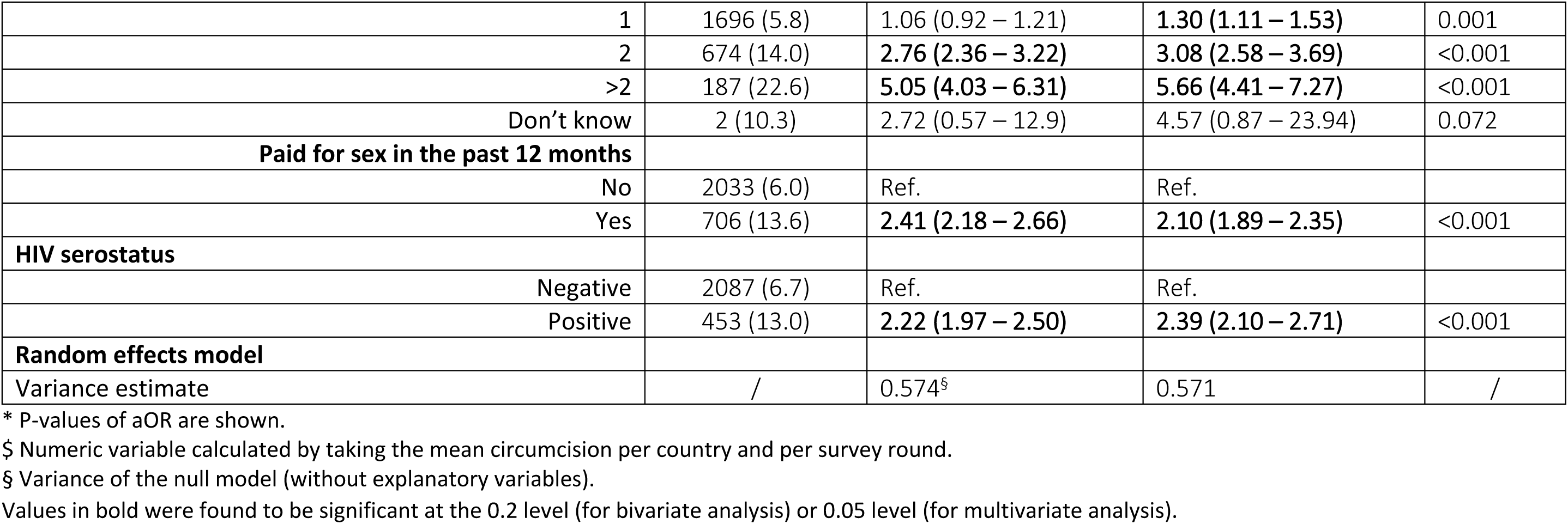
Results of the multilevel logistic regression analysis of the association between circumcision, other risk factors, and self-reported STIs (all survey countries and survey rounds pooled).

Supplementary file S3 shows the results of the sensitivity analysis using only self-reported STI diagnoses (excluding STI symptoms) as outcome variable. This analysis showed a harmful association between VMMC scale-up and self-reported STIs (aOR = 1.32; 95% CI 1.09 – 1.59; p=0.004). There was no association between an individual’s circumcision status and a self-reported STI diagnosis (aOR = 1.08; 95% CI 0.91 – 1.29; p=0.363). Supplementary file S4 shows the results of the sensitivity analysis based on post-2008 surveys looking at medical circumcision only (excluding traditional circumcision). After controlling for potential confounders, we observed a protective but non-significant association between and male medical circumcision status and self-reported STIs (aOR = 0.92; 95% CI 0.78 – 1.07; p=0.280).

## Discussion

This study aimed to investigate the association between male circumcision and self-reported STIs using representative, population-level data from Malawi, Rwanda, Zambia, and Zimbabwe; countries that scaled-up VMMC programmes since 2008. We found that VMMC scale-up at the population level was associated with a higher self-reported STI burden. At individual level, however, we found no evidence for an association between circumcision status and self-reported STIs. Although findings should be interpreted with caution, they add to the evidence base on the relationship between male circumcision and STIs other than HIV.

Our findings are consistent with, and further advance, the results of a previous study that found no association between individual circumcision status and self-reported STI symptoms using DHS data from 18 sub-Saharan African countries before VMMC scale-up (2003-2006) [24]. Notably, our multilevel analysis approach allowed to separate the population level exposure of VMMC scale-up (corresponding to increases in male circumcision coverage over time) from the individual circumcision status. Although we did not find any evidence of a protective association between male circumcision and STIs, and instead even report a harmful association with the population level exposure, it is important to understand these findings considering the methodological limitations of our before-after analysis. This includes, most importantly, the lack of a robust counterfactual scenario. It can, for instance, not entirely be excluded that male circumcision had a relative protective effect on STI acquisition in the context of an increasing background STI prevalence due to factors not comprehensively captured in DHS surveys but that coincided temporally with VMMC scale-up. Such secular epidemiological trends might also explain the discrepancy between the results of our two analyses (with main exposure at the population and individual level, respectively), and is further supported by our observation that STI prevalence increased over time among both circumcised and uncircumcised men. The observed increase in VMMC coverage over time may have been too small to curb this trend of rising STIs. To adequately control for such secular trends, one would need to conduct an experimental study with contemporaneous and randomly allocated exposure to either circumcision or no circumcision. However, given the proven prevention benefits of VMMC for HIV and some STIs, (cluster) randomised trials are likely to be unethical. Quasi-experimental designs (i.e. comparing groups with different levels of VMMC coverage) may offer a more suitable approach to advance the evidence on this topic.

Other potential, albeit less likely, explanations for the observed harmful association between VMMC scale-up and self-reported STIs could be enhanced STI testing and consultation rates resulting from the comprehensive sexual health promotion conducted as part of VMMC programmes. This expanded access to sexual health services could lead to increased STI diagnoses through improved detection of previously undiagnosed infections. However, the sensitivity analysis using only STI diagnoses as outcome (excluding STI symptoms) did not amplify this effect and showed similar results as the main analysis. Behavioural risk compensation among circumcised men represents another possible explanation. Although a 2017 study based on DHS data from 14 sub-Saharan African countries did not find any evidence of sexual risk compensation following VMMC scale-up, differences in (sexual) risk behaviour between circumcised and uncircumcised men that are not captured in DHS surveys cannot be entirely excluded in our study [25]. However, if risk compensation due to factors not adjusted for in our analysis were the primary mechanism driving increases in STIs, one would expect to also observe significant associations between STIs and circumcision status at the individual level, which we did not find.

Our second sensitivity analysis isolating the effect of medical (as opposed to traditional) male circumcision suggests a potential protective (but non-significant) effect on self-reported STIs. It has previously been shown that traditional circumcision often confers less protection against HIV and other STIs compared to medical circumcision, and may be harmful, due to partial removal of the foreskin, impaired healing, lack of counselling on sexual health, and sharing knives or blades during the procedure [26–29].

Other studies conducted in similar settings have provided more granular data on the relationship between male circumcision and biological STI diagnoses. For instance, findings from a household survey among 4640 women and 2850 men aged 15-49 years in Kwazulu-Natal, South Africa, showed a protective association between male circumcision and HSV-2, hepatitis B, and *mycoplasma genitalium*, but not syphilis, NG and CT [30]. Similarly, a study among 339 men aged 18-49 years at high risk of STIs in a South African mining community found a significantly higher prevalence of STIs, as diagnosed by syndromic assessment, among uncircumcised men compared to their circumcised men [31]. These findings are in line with earlier RCTs conducted in Kenya that showed no effect of male circumcision on the incidence of NG, CT and *trichomonas vaginalis* infections among men, while a protective association was found between male circumcision and HSV-2 in a hyperendemic South African setting using targeted maximum likelihood estimation [13,32]. Medical male circumcision may provide protection against ulcerative STIs (e.g. HSV-2 and syphilis) through the prevention of micro-abrasions that present a portal of entry and possible eruption site for these pathogens, while for urethritis-prone infections (e.g. NG and CT) the biological protection mechanism of circumcision is less clear and might be less pronounced [10]. In our study, the effect of male circumcision on different STIs cannot be distinguished due to the self-reported nature of our data, which likely excluded asymptomatic STIs while also misclassifying some cases as STI based on symptoms that resemble STIs (but which were not actual STIs). This could additionally explain the lack of an observed association in our analysis.

Weary of the potential measurement errors explained above, our study emphasises the high and rising STI burden in the sub-Saharan African region, particularly among sexually active boys and men, and despite increasing VMMC coverage. Our overall point prevalence estimate of the self-reported STIs in the four observed countries in 2015-2020 was higher than those reported among men aged 15-49 years in the African WHO region in 2016 for CT (4.0%; 95% CI 2.4–6.1%), NG (1.6%; 95% CI 0.9–2.6%) and Syphilis (1.6%; 95% CI 1.2–2.0%) [33]. It was also higher than the pooled prevalence of self-reported STIs among sexually active men in 10 Eastern African countries based on DHS data from 2011-2022 (5.22%; 95% CI 5.34–5.69%) [34]. Consistent with other studies, we found that young age (15-29 years old), high mobility, and lower education levels, were significantly associated with STIs [34–36]. Our study thus suggests that a focus on VMMC (including the services offered during VMMC) alone is insufficient to reduce the STI burden among boys and men at population level. However, VMMC clearly provides important benefits for the prevention of HIV and some specific STIs, including among women (e.g. HPV-associated cervical cancer). These insights, together with our finding that the observed circumcision coverage in the four countries remained below 50%, should encourage future studies to continue to focus on achieving a better integration of VMMC programmes with STI and other male-oriented services, alongside the development and implementation of male-friendly health services to reduce remaining barriers to STI services [37–39]. A valuable example includes a recent report of the effectiveness of a multi-component intervention to increase VMMC uptake among uncircumcised men in an STI clinic in Malawi [40].

## Conclusion

This study is among the first to use real-world, population-level data representative of the sexually active male population to explore the effect of VMMC scale-up on the prevalence of self-reported STIs in Eastern and Southern Africa. While we found no evidence of a protective association between male circumcision (scale-up) and self-reported STIs among adolescent boys and men (15-49 years), it cannot be entirely excluded that scaling-up VMMC programmes had a protective effect on the prevalence of specific STIs. Future studies with more rigorous designs and ability to differentiate between different STIs need to address this question more in-depth. However, while VMMC remains critical for HIV prevention, our results suggest that VMMC programmes alone are insufficient to address the broader STI burden among men. The high and rising STI burden in this population, together with the relatively low male circumcision rates observed in some settings, call for a better integration of VMMC programmes with comprehensive STI prevention and treatment services, alongside enhanced male-friendly health services.

## Data Availability

The datasets on which our analyses are based can be accessed at: https://dhsprogram.com/data/dataset/.

## Acknowledgments

We are grateful to the DHS Program for providing us with free access to the available datasets.

## Supporting materials

S1: Socio-demographics and sexual risk behaviour among men aged 15-49 in the four nationally representative pre-2008 population-based surveys.

S2: Socio-demographics and sexual risk behaviour among men aged 15-49 in the four nationally representative post-2008 population-based surveys.

S3: Results of the multilevel logistic regression analysis of the association between circumcision, other risk factors, and self-reported STI diagnoses (all survey countries and rounds pooled).

S4: Results of the multilevel logistic regression analysis of the association between medical circumcision, other risk factors, and self-reported STIs (only post-2008 surveys pooled).

